# Pan-Enterovirus Characterization Reveals Cryptic Circulation of Clinically Relevant Subtypes in Arizona Wastewater

**DOI:** 10.1101/2023.11.20.23297677

**Authors:** Daryn E. Erickson, Kyle M. Simmons, Zachary A. Barrand, Chase L. Ridenour, Paige B. Hawkinson, Lacey Lemke, Shayne P. Sellner, Breezy N. Brock, Alexis N. Rivas, Krystal Sheridan, Darrin Lemmer, Hayley D. Yaglom, W. Tanner Porter, Monique Belanger, Rachel M. Torrey, Aidan James R. Stills, Kiley McCormack, Matt Black, Wydale Holmes, Drew Rostain, Jeremy Mikus, Kimberly Sotelo, Emmen Haq, Reshma Neupane, Joli Weiss, Jasmine Johnson, Clancey Collins, Sarah Avalle, Chelsi White, Brandon J. Howard, Sara A. Maltinsky, Ryann N. Whealy, Nathaniel B. Gordon, Jason W. Sahl, Talima Pearson, Viacheslav Y. Fofanov, Tara Furstenau, Elizabeth M. Driebe, J. Gregory Caporaso, Jarrett Barber, Joel Terriquez, David M. Engelthaler, Crystal M. Hepp

## Abstract

**Background:** Most seasonally circulating enteroviruses result in asymptomatic or mildly symptomatic infections. In rare cases, however, infection with some subtypes can result in paralysis or death. Of the 300 subtypes known, only poliovirus is reportable, limiting our understanding of the distribution of other enteroviruses that can cause clinical disease.

**Objective:** The overarching objectives of this study were to: 1) describe the distribution of enteroviruses in Arizona during the late summer and fall of 2022, the time of year when they are thought to be most abundant, and 2) demonstrate the utility of viral pan-assay approaches for semi-agnostic discovery that can be followed up by more targeted assays and phylogenomics.

**Methods:** This study utilizes pooled nasal samples collected from school-aged children and long-term care facility residents, and wastewater from multiple locations in Arizona during July–October of 2022. We used PCR to amplify and sequence a region common to all enteroviruses, followed by species-level bioinformatic characterization using the QIIME 2 platform. For Enterovirus-D68 (EV-D68), detection was carried out using RT-qPCR, followed by confirmation using near-complete whole EV-D68 genome sequencing using a newly designed tiled amplicon approach.

**Results:** In the late summer and early fall of 2022, multiple enterovirus species were identified in Arizona wastewater, with Coxsackievirus A6, EV-D68, and Coxsackievirus A19 composing 86% of the characterized reads sequenced. While EV-D68 was not identified in pooled human nasal samples, and the only reported acute flaccid myelitis case in Arizona did not test positive for the virus, an in-depth analysis of EV-D68 in wastewater revealed that the virus was circulating from August through mid-October. A phylogenetic analysis on this relatively limited dataset revealed just a few importations into the state, with a single clade indicating local circulation.

**Significance:** This study further supports the utility of wastewater-based epidemiology to identify potential public health threats. Our further investigations into EV-D68 shows how these data might help inform healthcare diagnoses for children presenting with concerning neurological symptoms.

## IMPACT STATEMENT

Much of our response to infectious disease threats is reactive, even in the case of seasonally circulating viruses. Wastewater-based epidemiology has been successfully used for mitigation purposes throughout the SARS-CoV-2 pandemic, and has potential to be implemented for other pathogens that can cause severe infections. While vaccines and treatment beyond supportive care are not available for many infections, this method of surveillance can provide situational awareness to public health agencies, medical professionals, and the general population, possibly paving the way for earlier public health response.

## INTRODUCTION

The earliest evidence of enteroviruses dates back to 1580–1350 B.C [1]. A recovered Egyptian funerary stele produced during that timeframe depicts a priest with a shortened and withered leg, contemporarily believed to have been caused by poliovirus. Approximately 3,000 years later, in 1789, Dr. Michael Underwood described a “debility of the lower extremities in children,” which is taken as the first clinical description of poliovirus. In 1840, Dr. Jacob von Heine described “infantile spinal paralysis” and additionally theorized that the condition could be contagious [2]. The condition was suggested to be caused by a virus by Drs. Karl Landsteiner and Erwin Propper in 1908, and antibodies to the virus were discovered two years later [3]. Despite the long-standing interplay between humans and enteroviruses, and our success eradicating wild-type poliovirus, our understanding of the greater enterovirus genus is limited given their genetic and disease complexity.

The Enterovirus genus, within the Picornaviridae family, encompasses 15 viral species (Enterovirus A]–L and Rhinovirus A–C), of which seven are known to be infectious to humans (Enterovirus A–D and Rhinovirus A–C) [4]. While the prefix “entero” means “relating to the intestine”, the nearly 300 enterovirus subtypes composing the seven species infecting humans are known to be diverse in their clinical manifestations [5]. Poliovirus (i.e. a subtype of Enterovirus C) is the most well-known within the genus, given the hundreds of thousands of cases caused prior to the Global Polio Eradication Initiative in 1988. Polio case counts have dramatically decreased worldwide, with only a few countries reporting cases annually [6]. The virus commonly presents with flu-like symptoms in the case of symptomatic infection, including sore throat, fever, fatigue, nausea, headache, and stomach pain, and less frequently as paralysis [7]. Although wild poliovirus has been eliminated from the United States and most other countries, a case of acute flaccid myelitis (AFM) due to vaccine-derived poliovirus type 2 (VDPV2) was discovered in New York in July 2022. The investigation was accompanied by geographically proximal and genetically linked VDPV2-positive wastewater samples collected between May and December of 2022, underscoring the utility of wastewater for estimating duration and extent of viral shedding in a population [8].

Aside from poliovirus, there are other clinically-relevant enteroviruses that have been associated with varying presentations, including substantial outbreaks of hand foot and mouth disease (e.g. coxsackievirus A6 - CV-A6, [9]), neurologic disease (e.g. EV-D68, EV-A71, [10]), severe respiratory illness (e.g. EV-D68, [11][12]), and acute hemorrhagic conjunctivitis (e.g. CV-A24, [13]). Of these, EV-D68 has arguably been the most clinically important enterovirus within the United States over the past decade, first identified in 1962, and linked to a series of pediatric clusters of AFM in 2014 [14]. Since August of 2014, there have been 729 confirmed cases of AFM in the US [15], spiking in 2014, 2016, and 2018, thought to have been caused by EV-D68 [16] (Fig. 1). Although the biennial pattern observed in previous years indicated that AFM cases should have spiked in 2020 and 2022, seasonality of several respiratory viruses was disrupted during the initial years of the COVID-19 pandemic [17,18].

**Figure 1.**
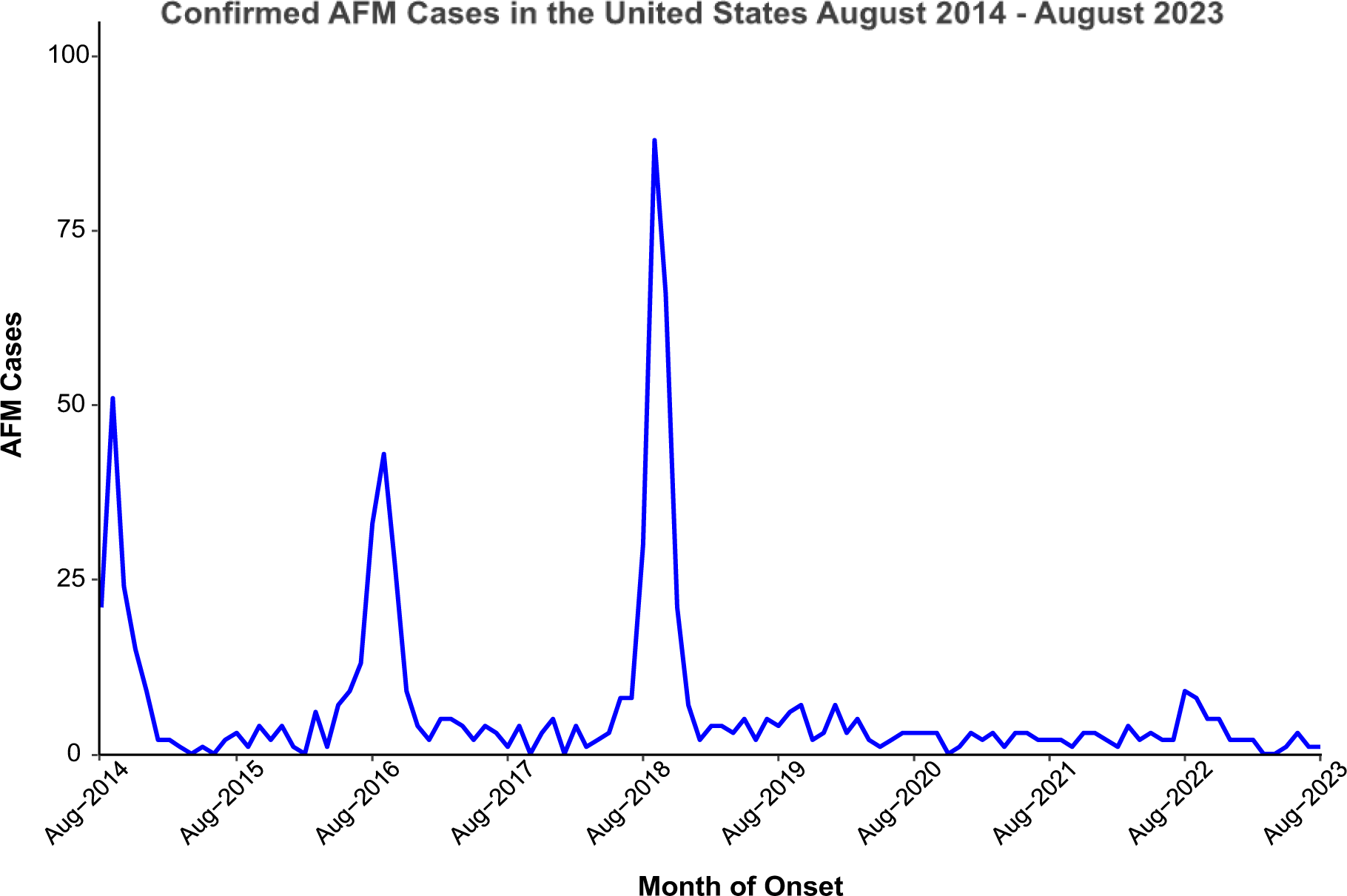
Acute Flaccid Myelitis cases in the United States over time. Initiation of surveillance by the Centers for Disease Control and Prevention was initiated in August 2014 and is shown here through August 2023. This figure was prepared using data from the CDC [15].

Most enteroviruses circulate undetected due to a predominance of asymptomatic or mildly symptomatic infections and are only further investigated as a reactive response to syndromic surveillance, or as part of a disease outbreak response. In an effort to better inform public health partners of circulating threats in the absence of or prior to being triggered by syndromic surveillance evidence, our team has been conducting respiratory virus surveillance in populations of school-aged children in Coconino County, Arizona and long-term care facility residents in Maricopa County, Arizona, and wastewater-based epidemiology for the same pathogens at congregate living sites and wastewater treatment plants (WWTPs) in the City of Flagstaff, and City of Tempe Biointel Basins. Here, we present our efforts to deploy a pan-enterovirus assay coupled with high throughput sequencing, and a novel use of the QIIME 2 bioinformatics platform [19] for the purposes of viral discovery and classification followed by near-whole viral genome sequencing of EV-D68.

## METHODS

### Sample and Clinical Data Collection

#### Wastewater

Raw wastewater samples used for this project were collected from two City of Flagstaff wastewater treatment plants from April 27, 2022, through October 26, 2022 (n=30), eleven Flagstaff congregate living settings from September 7 through October 26, 2022 (n=52), and eleven City of Tempe Biointel Sampling Basins (i.e. metering, lift, and wastewater sampling stations) from August 5, 2022 through October 26, 2022 (n=61). At the wastewater treatment plants and Biointel Sampling Basins, 24-hour composite samples were collected (125mL every 30 minutes) as part of ongoing operations. From each composite sample, we collected 80mL split evenly into two 50mL conicals. Samples from the eleven congregate living settings were collected from proximal manholes using an approach inspired by the Moore Swab. Briefly, Moore described their technique as “taking a piece of gauze about four feet in length and six inches wide, folding it into a pad of eight thicknesses and attaching it firmly (usually with fishing wire) by one end to a long piece of string. The gauze was immersed into the flowing sewage, the string attached suitably just under the manhole cover, and the gauze left in position for 48 hours [20], [21].” Instead of gauze, we immerse ultra-sized Tampax tampons attached to Rexlace plastic craft lace tied to a plastic hook glued with Gorilla Glue Construction Adhesive to the inside of the manhole. After 48 hours, tampons were pulled from the wastewater flow and placed in a plastic 125 mL jar with 20mL of molecular grade water and shaken vigorously.

#### Nasal Swabs

We retrospectively tested 809 nasal swabs collected from 146 school-aged children in Coconino County and 92 long-term care facility residents in Maricopa County, biweekly, from August through October of 2022. Each individual swabbed their own nose for 15 seconds per nostril with a micro flocked swab followed by storage in liquid Amies solution at 4°C prior to processing. All school-aged children attended schools within the tested sewersheds in the City of Flagstaff, while residents of long-term care facilities live adjacent to the sewersheds tested in the City of Tempe.

#### Clinical Data

Flagstaff Medical Center serves the 70,000 citizens of Flagstaff and the greater Northern Arizona region. The hospital’s microbiology lab provides standard of care testing for all patient samples. Electronic medical record data was queried to identify all instances of hospital testing for enteroviruses/rhinoviruses from 07/04/2022-12/31/2022 aggregated by week of sampling.

### Wastewater Concentration and Extraction

Immediately following collection, samples were concentrated using the Environ Water RNA Kit (Zymo Research) following manufacturer protocol for viral enrichment and sample homogenization. Promptly after sample homogenization, samples were extracted using the Quick-DNA/RNA Pathogen Miniprep Kit (Zymo Research), including the optional Proteinase K treatment, following manufacturer protocol. RNA was eluted into 100µL of DNase/RNase free water pre-warmed to 37°C.

### Wastewater RT-qPCR and cDNA Synthesis

Post extraction, all samples were also treated using Invitrogen’s ezDNAse reagent. The 10μL ezDNase reactions contained 8μL RNA input and were incubated for minutes at 37°C following manufacturer recommendations. The full volume of each DNase-treated sample was reverse transcribed using the SuperScript^TM^ IV First Strand Synthesis System (Invitrogen) following manufacturer recommendations with modified thermal cycling conditions. Initial primer annealing with random hexamers was performed with maximum RNA input and incubated at 70°C for 7 minutes. Following primer annealing, reverse transcription of RNA was performed by cycling at 23°C for 10 minutes, 50°C for 45 minutes, 55°C for 15 minutes, and 80°C for 10 minutes. All synthesized cDNA was treated with the included RNase H to remove RNA in the RNA/cDNA hybrids that formed during reverse transcription. cDNA was stored at -20°C awaiting further analyses.

Following concentration and extraction of wastewater, samples were tested for the presence of or absence of EV-D68 using real-time quantitative PCR (RT-qPCR) methods, using the CDC2022 EV-D68-specific primers and probe, Forward: AN993, Reverse: AN995, and Probe: AN992 [47]. Each 20µL RT-qPCR reaction contained Luna® Probe One-Step RT-qPCR 4X Mix with UDG (New England BioLabs) at final concentration of 1X, and forward primer, reverse primer, and probe all with the final concentration of 0.5µM. All reactions contained 5µL of RNA template input. Thermal cycling conditions were as follows: carryover prevention at 25°C for 30 s, reverse transcription at 55°C for 10 min, activation and denaturation at 95°C for 1 min, and 40 cycles of 95°C for 10 s, 55°C for 30 s. Viral load in positive samples was calculated using a synthetic DNA control, designed in-house, of known concentration. Importantly, the CDC2022 assay was validated by the CDC using a 7500 Fast Real-Time PCR System (Applied Biosystems) with qScript™ XLT One-Step RT-qPCR ToughMix® (Quanta Biosciences), and may have performed differently given different equipment and reagents used here. In addition to the CDC2022 RT-qPCR assay, we also selected a subset of samples to be tested with a new RT-qPCR that our team developed, termed the “D68-Detect” assay, targeting a 108bp fragment of the 2C gene due to its higher level of conservation. Each 10µL RT-qPCR reaction contained Luna® Probe One-Step RT-qPCR 4X Mix with UDG at final concentration of 1X, D68-Detect-F sense primer (T<u>M</u>CATGGCTCTCCAGGAACT) and D68-Detect-R antisense primer (CTTAGGGTCTGGGGGCA<u>R</u>GG) at a final concentration of 0.4µM, and D68-Detect-P probe (FAM-TGGCCTCAAATTTAATTGCCAGGGC) at a final concentration of 0.2µM, covering positions 4,440-4,546. All reactions contained 2µL of RNA template input. Thermal cycling conditions were the same as those used with the CDC2022 assay. Viral load in positive samples was calculated using an EV-D68 positive control isolated in 2018 from BEI Resources (NR-52357) in which concentration was determined using Droplet Digital PCR (ddPCR). All RT-qPCR reactions for both assays were carried out on a QuantStudio^TM^ 7 Flex Real-Time PCR System (Applied Biosystems).

### DDPCR for positive control quantification

The EV-D68 isolate from BEI Resources (NR-52357) underwent cDNA synthesis using New England BioLabs Luna® Script RT Supermix kit following manufacturer recommendations. A 10-fold serial dilution was performed on the newly synthesized cDNA down to a 1:10,000,000 dilution. Droplet Digital PCR was performed on the dilution series using the newly designed RT-qPCR assay described above on a BioRad QX200 AutoDG Droplet Digital PCR System, however, the first dilution was omitted to avoid 100% positive droplets in the reaction. The 22µL ddPCR reactions consisted of 2X ddPCR Supermix for Probes (No dUTP), forward and reverse primers, and probe at final concentrations of 1X, 0.9µM, and 0.25µM respectively. Following droplet generation on the automated droplet generator, samples were cycled on a BioRad C1000 Touch Thermal Cycler with 96-Deep Well Reaction Module using manufacturer recommended cycling conditions with slight modification. Reactions were cycled for a total of 50 cycles and an annealing temperature of 55°C. Following cycling, droplets were read using a Direct Quantification experiment on a BioRad QX200 Droplet Reader. Thresholds were manually set for all samples using amplitudes of the Reverse Transcription and ddPCR No Template Controls to normalize positive droplet thresholding across samples (Fig. S2a). Final concentrations were back-calculated across each dilution and an average concentration was determined (Supplemental Table 1). Dilutions with too few or too many positive droplets were omitted from the average concentration calculation (Table S1, Fig. S2b).

### Nasal Swab Testing

Liquid Amies solution from 2–6 swabs was pooled together to a final volume of 50µL and digested following Workflow 1 from the SalivaDirect^TM^ protocol [22]. Digested pools were then tested for the presence or absence of EV-D68 using the same RT-qPCR methods described previously.

### PCR Amplification and Sequencing

#### Pan-Enterovirus Assay

Synthesized cDNA from all samples was amplified using previously designed pan-enterovirus primers targeting a conserved 440bp’ UTR region of the Enterovirus genome [23], with attached universal tails [24]. Each 20uL PCR reaction contained Invitrogen PCR Buffer, Invitrogen MgCl_2_, Invitrogen dNTPs, Invitrogen PlatTaq, and primers at final concentrations of 1X, 2mM, 0.2mM, 0.1U/µL, 0.375µM and respectively. Additionally, the reactions contained 2µL of cDNA template input. Thermal cycling conditions were as follows: initial denaturation at 95°C for 1 min, 35 cycles of 95°C for 15 s, 60°C for 15 s, 72°C for 30 s, and a 7-minute final extension at 72°C. Amplified product was cleaned using 0.8X Agencourt AMPure XP beads (Beckman Coulter).

#### EV-D68 Tiled Amplicon Assay

Primal Scheme [25] was used to develop a multiplex of tiled EV-D68-specific primers. A total of 23 primer pairs with attached Illumina-compatible universal tails were designed, covering genome positions 60–7308 of a contemporary EV-D68 genome (OP267522.1), amplifying regions averaging 356bp. The primer set was assessed using three positive controls from the Biodefense and Emerging Infections Research Resources Repository (BEI Resources): NR-49135 (accession MH708882) [63], NR-52357 (accession MN246009), and NR-55939 (isolate sequence not yet available). Synthesized cDNA from samples identified positive for EV-D68 with RT-qPCR with Ct values less than or equal to 35 were amplified using these newly designed primers (Table S4), with the primers separated out into two pools.

#### Library Preparation and Sequencing

A second PCR using universal tail-specific primers was performed to add the Illumina specific indexes [24]. Each uL indexing PCR reaction consisted of 1. μL of X Kapa HiFi HotStart Ready Mix (Kapa Biosystems) for a final 1X concentration, 400 nM of each forward and reverse index primer, and 4, 6, or 8 µL of the cleaned amplified Pan-enterovirus product. Reactions were cycled as follows: 98°C for 2 min, 6 cycles of 98°C for 30 s, 60°C for 20 s, and 72°C for 30 s, and a final extension at 72°C for 5 min. Indexed samples were cleaned using 0.8X Agencourt AMPure XP beads (Beckman Coulter). Cleaned, indexed product was quantified using the Kapa Library Quantification kit (Kapa Biosystems) on an Applied Biosystems QuantStudio 7Flex System. The samples were then pooled equal molar and final sample pools were sequenced on one of three of the following Illumina platforms: MiSeq (using a v2 500 cycle kit), NextSeq 1000 (using a P1 600 cycle kit), NovaSeq 6000 (using an SP v1.5 500 cycle kit).

### Statistical Analyses

To investigate the leading predictive ability of viral load for test positivity of viral infection, a low-rank penalized regression spline was fit to each of the viral load (average copies per mL) and viral test positivity count data sets as functions of time [26]. These are semi-parametric models whose smoothness and complexity are determined via data-based out-of-sample prediction optimality criteria. Then each model was used to obtain daily interpolated predicted values— viral load and test probability of positivity—over a range of time, and the predicted (log odds of) test probabilities of positivity where regressed on the predicted (log) viral loads for a range of time lags for which test results are expected to lag loads, or, conversely, load values are expected to lead test results.

### Bioinformatic Processing

To characterize all enteroviruses sequenced using the pan-assay, we took a marker gene-based approach as has been well-documented for microbiome analyses using QIIME 2 [19]. This approach involves the development of an appropriate reference database, and classifier training, and classifying paired-end sequencing reads with the trained classifier. For samples where EV-D68 was detected, additional near-whole genome sequencing was carried out followed by phylogenetic reconstruction.

#### Curating a Picornaviridae Reference Dataset

Reference sequences were downloaded from the NCBI nucleotide database using QIIME 2 2022.11 [19] plugin RESCRIPt [27]. Using the method get-ncbi-data with the NCBI search query of “Picornaviridae[ORGANISM] AND 5000:10000000[SLEN]”, which finds all sequences for the virus family Picornaviridae with sequence length over 5,000 base pairs [28][29]. The search resulted in 15,473 downloaded sequences. The downloaded sequences were trimmed to the target region with the q2-feature-classifier plugin’s [30] extract-reads method with the forward and reverse pan-enterovirus primers as input. If the primers had more than three mismatches, the sequence was removed from the dataset. After extracting the target region, 8,635 sequences remained. To ensure the reference database did not contain any identical sequences, we used RESCRIPt’s dereplicate method, a wrapper for VSEARCH [31] methods, to remove duplicated sequences. After dereplication, the final reference dataset and taxonomy files contained 5,039 sequences and taxon, respectively.

#### Training Naïve Bayes Classifier

The naïve-Bayes classifier was trained using RESCRIPt’s evaluate-fit-classifier [27], a wrapper over scikit-learn’s [32][33] built-in naïve-Bayes classification methods. We trained the multinomial naïve-Bayes classifier using the reference dataset from the previous section with default parameters. The classifier was validated by k-fold cross-validation [30,34].

#### Preparing and Classifying Sequences with Trained Naïve-Bayes Classifier

The 143 wastewater sample reads, from 30 City of Flagstaff, 61 City of Tempe, and 52 congregate living sites, were trimmed using QIIME’s Cutadapt [35] plugin to remove primer and adapter sequences. After trimming, the reads were clustered into amplicon sequence variants (ASVs) using the QIIME DADA plugin’s denoise-paired method [36], resulting in 901 unique ASVs. The ASVs were taxonomically annotated using the classifier trained in the previous section using QIIME 2 feature-classifier [30] classify-sklearn [32,33].

#### EV-D68 Consensus Sequence Generation

Virus genome consensus sequences were built using the Amplicon Sequencing Analysis Pipeline (ASAP) [37][38]. First, reads were adapter-trimmed using bbduk [39], and mapped to an EV-D68 reference genome (OP267522.1, [40]) with bwa mem [41] using local alignment with soft-clipping. BAM alignment files were then processed to generate the consensus sequence and statistics on the quality of the assembly by the following: 1) Individual basecalls with a quality score below 20 were discarded. 2) Remaining basecalls at each position were tallied. 3) If coverage ≥10X and ≥80% of the read basecalls agreed, a consensus basecall was made. 4) If either of these parameters were not met, an ‘N’ consensus call was made.) Deletions within reads, as called during the alignment, were left out of the assembly, while gaps in coverage (usually the result of a missing amplicon) were denoted by lowercase ‘n’s. This method has previously been used to generate SARS-CoV-2 consensus genomes [42,43]. Statistics reported for each sample included: total reads, number of reads aligned to reference, percent of reads aligned to reference, coverage breadth, average depth, and any SNPs and INDELs found in ≥10% of the reads at that position.

#### EV-D68 Phylogenetic Analysis

Public genomes were selected from the NCBI Virus database based on genome completeness (at least 7,000 bases in length) and metadata availability, where country of origin was included at a minimum. Additionally, genomes from non-human hosts and duplicates from the same sample were removed. Consensus genomes built using ASAP along with 935 publicly available EV-D68 genomes were aligned using Multiple Alignment using Fast Fourier Transform (MAFFT) [44]. Model selection and maximum likelihood reconstruction was carried out using IQTree v1.6.12 [45], with the ModelFinder [46] identifying GTR+F+R5 as the best fit model. The resulting consensus tree file, including the associated bootstrap values, was then input into iTOL [47] for visualization. Genome clade determination (A, B1, B2, B3, C, D) was based on a previous study [48]. A sub-tree was annotated from the same dataset to better visualize the genomes obtained during this study (Fig. 4) using FigTree [49].

## RESULTS

### Enterovirus Discovery

From April 27 to October 26, 2022, we screened a total of 177 wastewater samples from congregate living sites, wastewater treatment plants, and Biointel Sampling Basins in Coconino and Maricopa counties, using previously published pan-enterovirus primers (Primer 1 and Primer 3 of the ‘-NCR) [23]. Of those samples, 143 demonstrated banding indicative of a 440bp amplification product when agarose gel electrophoresis or fragment analysis on a tapestation was performed and were moved forward for sequencing. Of these, four samples produced no enterovirus-related reads (Fig. 2, black columns), while amplification products from the remaining 139 samples resulted in 35 to 83,113 reads, with a mean of 9,926 and a median of 1,279 reads (Fig. 2 and Table S2).

**Figure 2.**
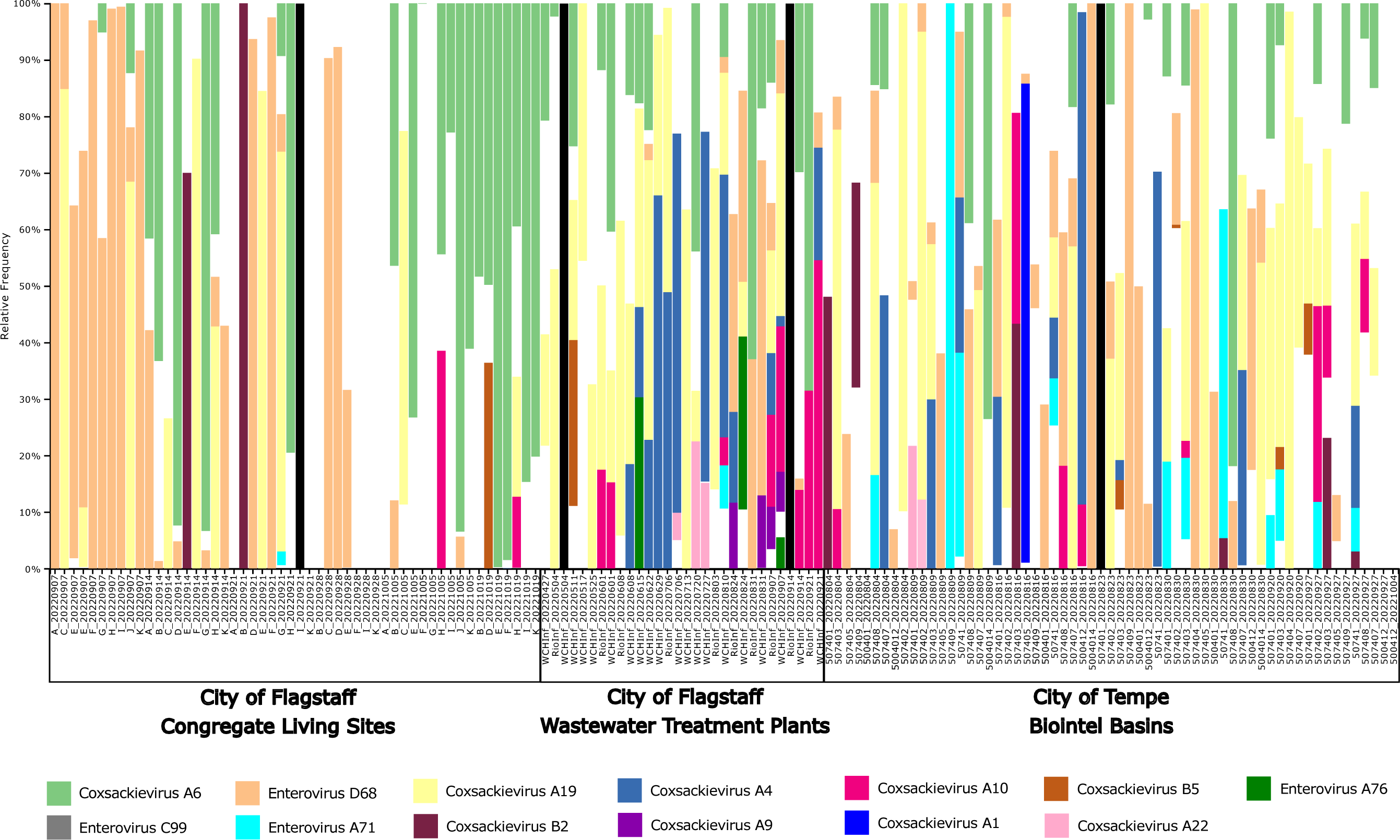
Taxonomic composition bar plot illustrating the species-level composition of enteroviruses in wastewater samples. Enterovirus subspecies that had at least 1% of the total reads classified as that subspecies are shown in this plot. A more detailed summary of the read classification per sample can be found in Table S2. Samples were sorted by Location Type and Collection Date. Columns showing only black bars indicate samples where no sequencing reads were produced, and white indicates uncharacterized reads

Sequencing reads from wastewater samples collected during this project were either classified as one of different enteroviruses, or “uncharacterized”. The latter indicates that the combination of the selected reference database coupled with the QIIME 2-based approach could not classify the reads because 1) they were not from enteroviruses (e.g. non-specific amplification), 2) the classification method was too conservative (i.e. did not allow for enough mismatches), 3) the reference database included too much overlap, preventing conclusive subspecies calls, or 4) that novel enteroviruses were present in the collected samples. Enteroviruses that accounted for at least 1% of reads across all sites using the pan-enterovirus assay, in order of greatest to least number of reads, were CV-A6, 34%, EV-D68 (17%), CV-A19 (7%), CV-A4 (2%), and CV-A10 (2%). However, CV-A4 was not present at the congregate living sites. CV-B5, EV-C99, EV-A71, CV-A9, CV-A1, and EV-A76 were limited to Biointel Sampling Basins and wastewater treatment plants. Notably, we also detected Enterovirus G at Northern Arizona (Flagstaff) wastewater treatment plants. This enterovirus species is generally associated with domestic and wild pigs but has been recently detected in human wastewater sources in the southern part of the state [50].

### EV-D68 Viral Load in Two Arizona Communities

The pan-enterovirus assay revealed the presence of EV-D68 in 65 samples (Fig. 2, Table S2). We further investigated EV-D68 viral loads in wastewater using an RT-qPCR assay recently published by the CDC [51], as well as the D68-Detect assay developed as part of this project. The first sample found to have EV-D68 reads using the pan-enterovirus assay dated to June 22, 2022 (n=20 reads), however neither RT-qPCR assay resulted in detection. Pan-enterovirus sequencing reads from this sample also revealed the higher relative abundance of CV-A4, CV-A6, and CV-A19, which are more likely to be responsible for the agarose gel electrophoresis banding that prompted further processing of this sample for pan-enterovirus sequencing. Using both the RT-qPCR and pan-enterovirus approaches, EV-D68 was not detected again until August 10, 2022, at the City of Flagstaff’s Wildcat Hill wastewater treatment plant. An additional 114 samples were found to be RT-qPCR positive in the late summer and early fall of 2022, where the viral load in both Tempe and Flagstaff peaked in late August and declined throughout September (Fig. 3a-d). In both cities, the D68-Detect assay estimated a higher viral load compared to the CDC2022 assay which could be due to differences in optimization and validation methods (e.g. positive control(s) used, RT-qPCR platform). The last RT-qPCR positive wastewater samples from Tempe and Flagstaff were collected October 11 and October 19, 2022, respectively. In contrast to wastewater samples, none of the 382 nasal samples from school-age children in Coconino County or long-term care facility patients in Maricopa County were positive for EV-D68.

**Figure 3.**
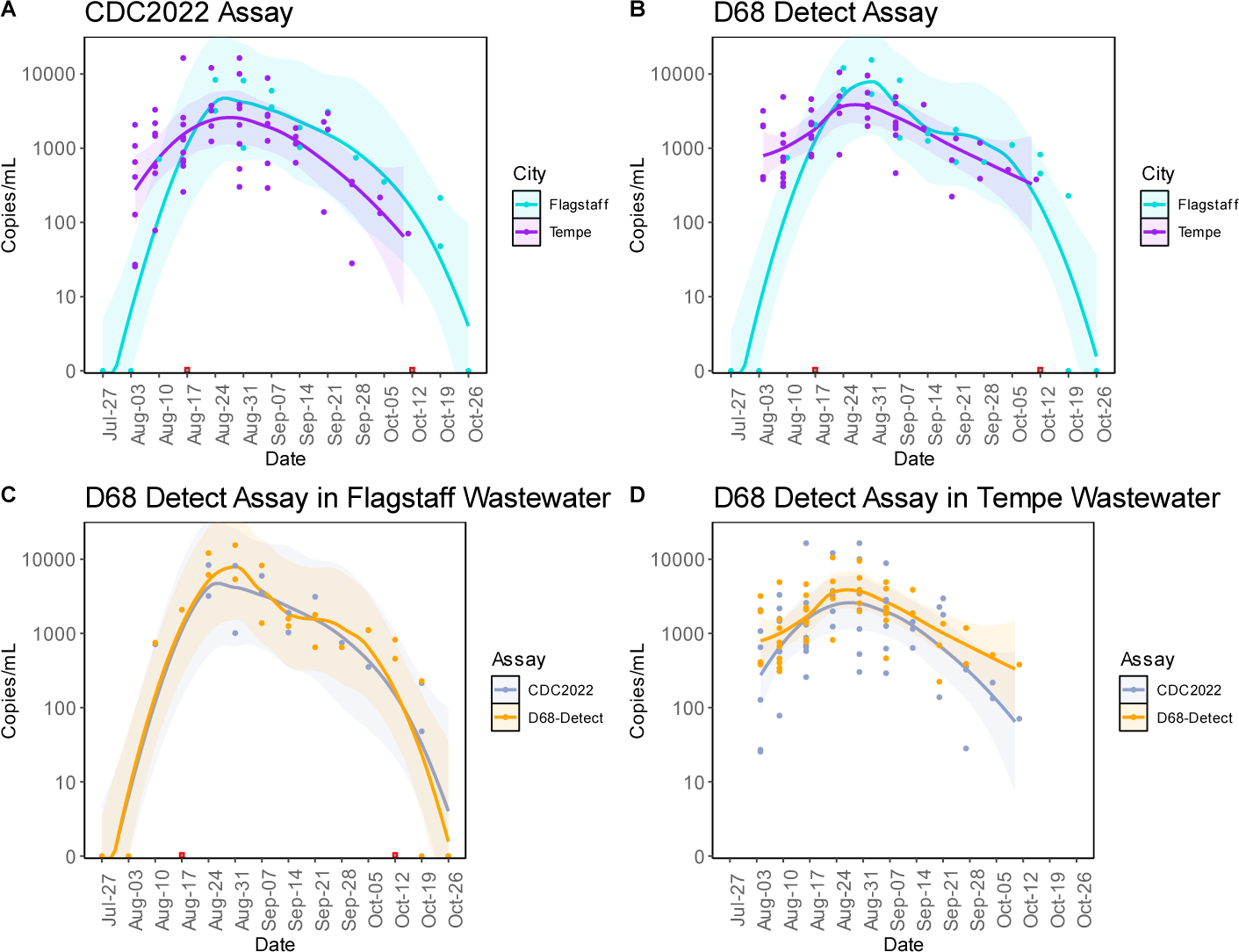
Trends of EV-D68 in City of Flagstaff and City of Tempe wastewater. EV-D68 viral loads in City of Flagstaff WWTPs and Tempe Biointel Sampling Basins between July and October 2022. Viral load in both cities was assessed using two different RT-qPCR assays: CDC2022 (A) and D68-Detect (B) with City of Flagstaff in teal and City of Tempe in purple. Assay comparisons for each city can be seen in plots C (Flagstaff) and D (Tempe) with CDC2022 in gray and D68-Detect in orange. A trend line was fitted to each set of data points on each plot coupled with a 99% confidence interval around each trend line. Data points in red boxes represent true zeros in viral load detection that were removed from the trend lines and confidence intervals then reintroduced into the plots.

A direct comparison of EV-D68 load in City of Flagstaff wastewater versus total enterovirus/rhinovirus percent positivity at Flagstaff Medical Center indicates that wastewater trends led hospital trends by 3 days. Both the Akaike information criterion and generalized cross-validation were optimized with this lag, with change in wastewater explaining 98.6% of the deviance in hospital positivity rates. However, the wastewater date of collection used in this analysis was the date the samples were collected, while the hospital date is the last day in the reporting week, and includes cases reported throughout the entire week. Therefore, the three-day lag between EV-D68 load in wastewater and enterovirus/rhinovirus percent positivity at the hospital indicates essentially no difference between the two surveillance methods.

### Efficacy of a New Tiled Amplicon Sequencing Scheme

A multiplex of tiled EV-D68-specific primers covering genome positions 60–7308 was designed as part of this study for targeted sequencing of 31 contemporary EV-D68 genomes from Arizona wastewater. Efficacy of the multiplex was assessed on three different positive controls (PTCs) from BEI Resources from 2014 (NR-49135), 2018(NR-52537), and 2020(NR-55939). Breadth of coverage ranged from 86-100% within the 60-7308 position range, with depth of at least 50x (Fig.5). Each PTC was sequenced on an Illumina NextSeq 1000 system using a P1 600 cycle kit and results were as follows: 2014 PTC received 10,221,430 paired-end reads (3.93% of lane), 2018 PTC received 15,921,338 paired-end reads (6.12% of lane), 2020 PTC received 18,979,870 paired-end reads (7.29% of lane). Phylogenetic analysis of the controls revealed each control clustered in three different subclades (B1, B3, and D respectively). The 2018 control performed the best with just 1.90% of the genome receiving less than 50X coverage and more than 92% of the genome receiving greater than 10,000X coverage. The 2020 control performed the worst of the three with 13.43% of the genome receiving less than 50X coverage and only 67.75% receiving greater than 10,000X coverage. Efficacy of the multiplex on the 2014 control was comparable to the 2018 control with less than 3% of the genome receiving less than 50X coverage and 84.54% receiving greater than 10,000X coverage. Through our validation of the new tiled primer multiplex using PTCs, we were able to assess the panel’s efficacy on diverse EV-D68 genomes. The pairwise percent identity between the PTCs and 2021 reference genome (OP267522.1) used to design the multiplex can be seen in Table 1. The contemporary reference genome and PTCs used represent three different subclades (D, B1, B3) and percent identity between any of these genomes ranged between 85.03-95.87%.

**Figure 4.**
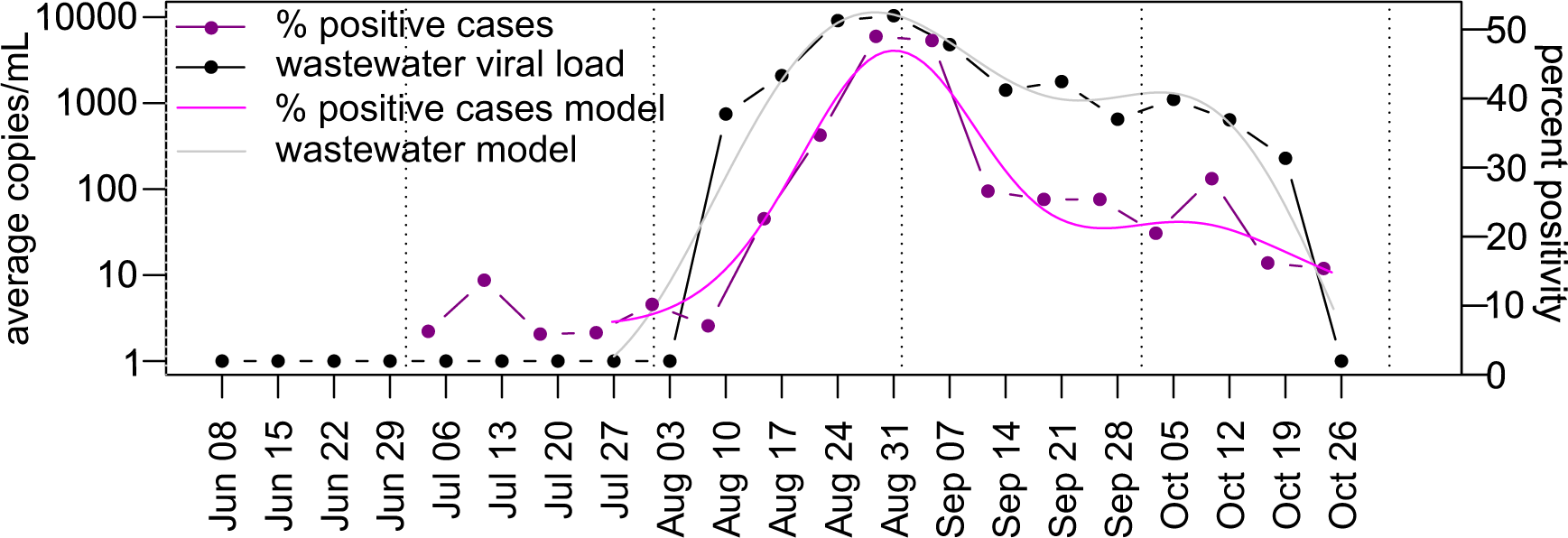
Trends of enteroviruses/rhinoviruses in clinical samples. Percent positivity at Flagstaff Medical Center between June and October 2022 compared to EV-D68 viral load in two City of Flagstaff wastewater treatment plants. Viral load was assessed with the D68-Detect assay. Dotted vertical lines indicate the 1st of each month.

**Figure 5.**
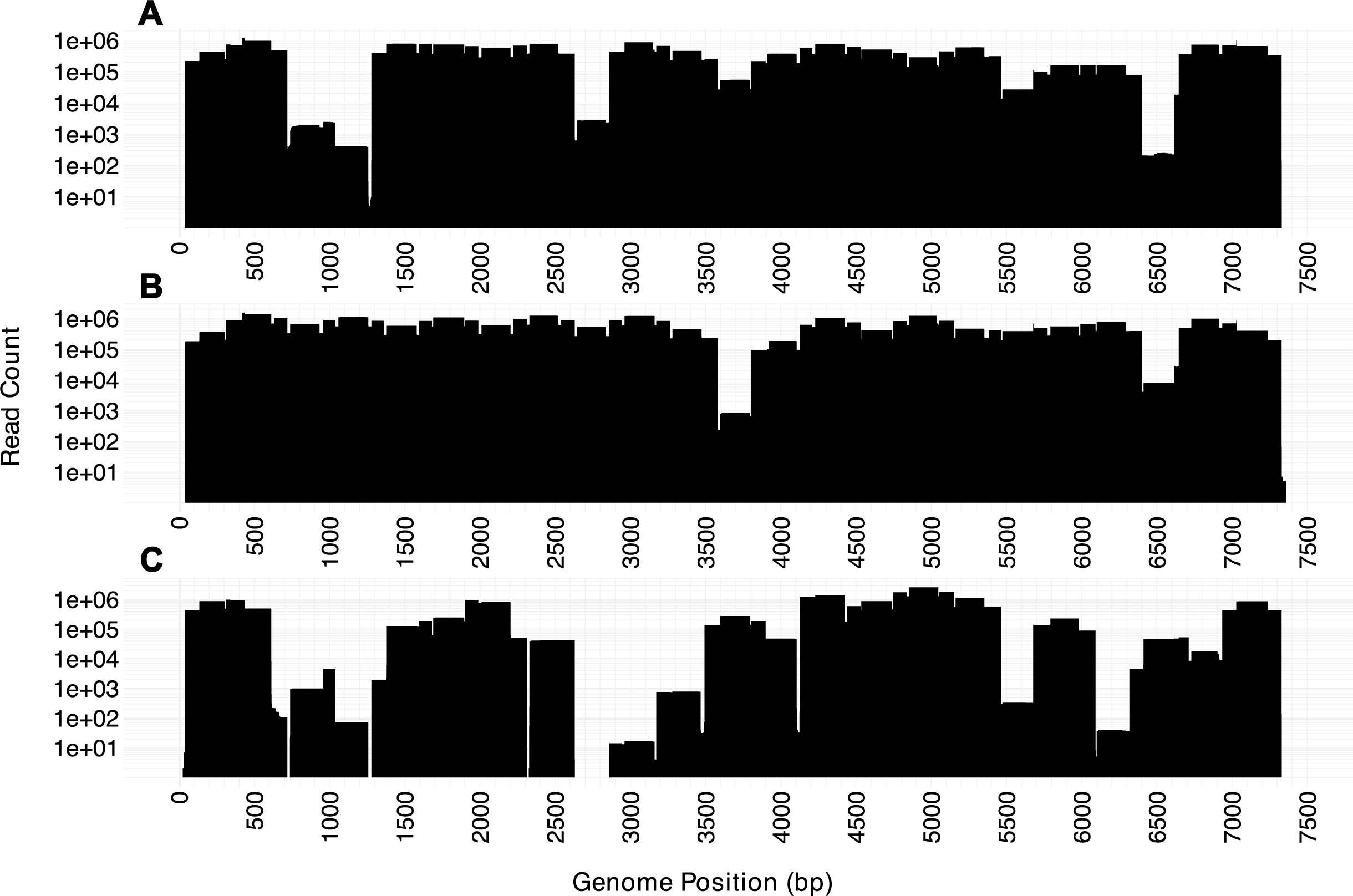
Genome coverage using the newly developed tiled amplicon design. The number of sequencing reads, displayed on the y-axis (log-scale), across all positions of each genome are shown. A) NR-49135 (accession MH708882), B) NR-52357 (accession MN246009), and C) NR-55939.

**Figure 6.**
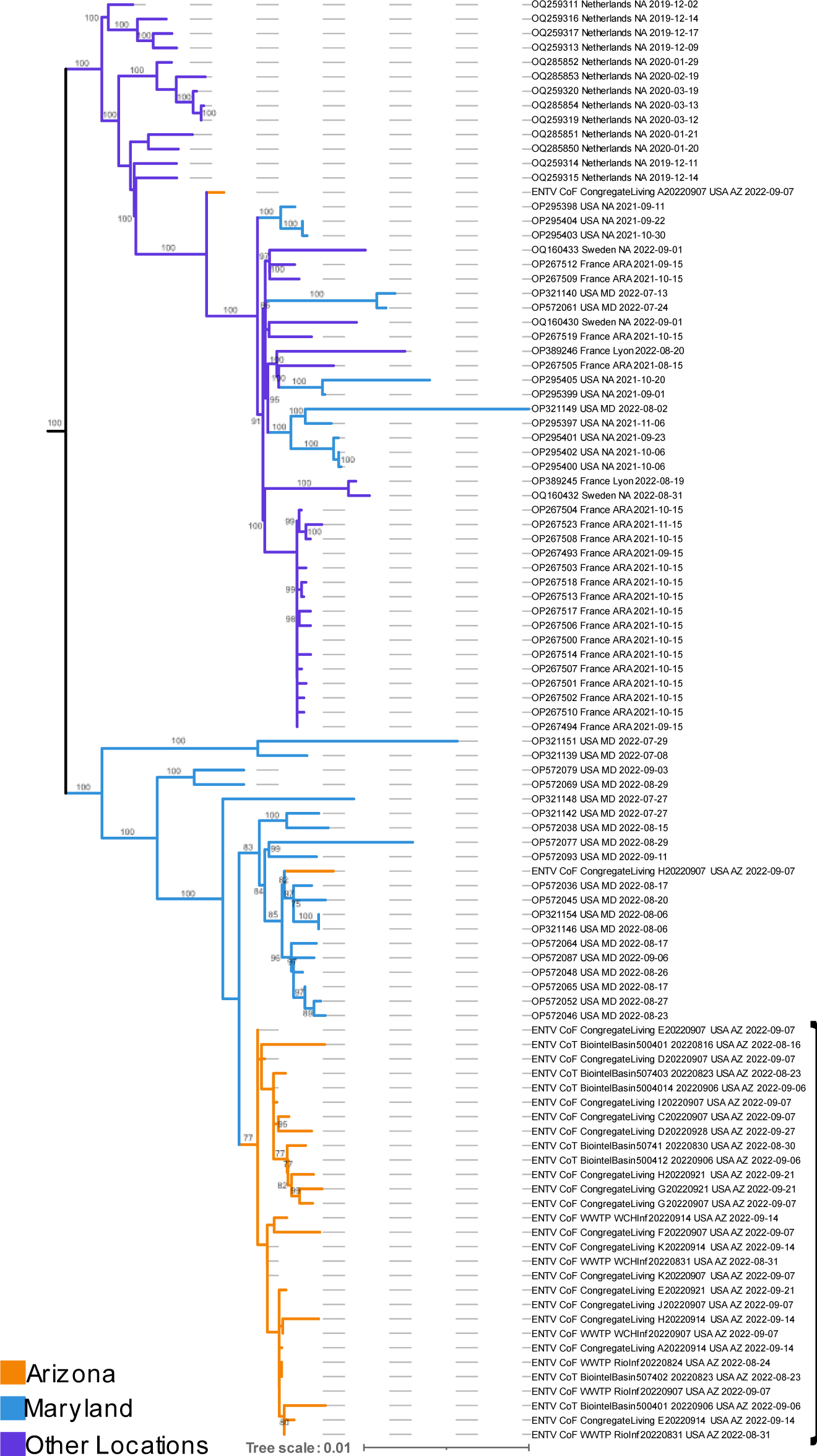
Subset of global EV-D68 maximum likelihood phylogeny. 69 of 935 publicly available EV-D68 genomes from NCBI and 31 new AZ genomes generated as part of this study. Bootstrap values indicating cluster support are on branches. Full phylogeny can be visualized in Supplementary Figure 1.

**Table 1.** Percent identity matrix between three EV-D68 positive controls and contemporary reference genome using the newly developed tiled amplicon sequencing panel. NR49135 (Isolated 2014, Subclade B1), NR52357 (Isolated 2018, Subclade B3), NR55939 (Isolated 2020, Subclade D), and the reference genome OP267522.1 used to develop the tiled amplicon sequencing scheme (Isolated 2021, Subclade B3).

|  | NR-55939 | NR-49135 | NR-52357 | OP267522.1 |
| --- | --- | --- | --- | --- |
| NR-55939 | 100% | 85.97% | 85.73% | 85.03% |
| NR-49135 | 85.97% | 100% | 94.84% | 93.71% |
| NR-52357 | 85.73% | 94.84% | 100% | 95.87% |
| OP267522.1 | 85.03% | 93.71% | 95.87% | 100% |

### Application of a New Tiled Amplicon Sequencing Scheme on Wastewater Samples

Once the tiled amplicon primer set was assessed on positive controls, we applied the same protocol to 31 wastewater samples that had tested positive for EV-D68 using both RT-qPCR assays. The total breadth of coverage ranged from 61.36-97.47%, with at least 10x coverage, when using OP267522.1 as a reference genome. While Illumina platforms were used to sequence genomes from all 31 samples, those sequenced on a NovaSeq 6000 (SP v1.5 500 cycle kit) received more paired end reads (1,556,926-8,623,394) than samples sequenced on a MiSeq using a v2 500 cycle kit (22,084-311,666). Correspondingly, EV-D68 genomes sequenced on the NovaSeq 6000 had a median breadth of coverage of 92.21%, versus 76.55% for the MiSeq, not indicating a difference between the platforms, but rather the greater number of reads allocated per sample. The percentage of sequenced reads mapping to EV-D68 ranged between 0.7-66.2%, indicating that the tiled amplicon primers generated a substantial amount of non-specific binding and amplification, or that residual DNA from the starting sample was carried through the entire process.

### Phylogenetic Analysis of EV-D68 from Arizona within a Global Context

A global maximum likelihood phylogenetic reconstruction was carried out to provide context around dispersal of EV-D68 into and within Arizona. All 31 Arizona wastewater genomes were part of the B3 subclade (Fig. S1). Of those, 30 were nested within genomes collected in Maryland, USA, overlapping the same time frame — July to September of 2022 (Fig. 4, bootstrap support of 100). Twenty-nine clustered monophyletically within the Maryland genomes, although not by city, location type, or date of collection. Importantly, bootstrap support for the cluster of 29 Arizona wastewater genomes was lower than for clusters primarily composed of clinical genomes (Fig. 4). This is likely the result of a larger number of uncalled sites (N’s) that arose either due to lower genome coverage or mixed nucleotide calls (Table S3), both common for viruses sequenced from wastewater. Total paired-end reads and EV-D68 mapped reads for all 31 genomes and three PTCs sequenced as part of this study can be found in Table S5.

## DISCUSSION

We initiated EV-D68 wastewater-based surveillance in November of 2021 with a now-deprecated RT-qPCR assay that we had originally designed for routine use. This initial assay never detected EV-D68 in human or wastewater samples but was more sensitive than the CDC2015 assay when used on controls dated 2018 and earlier. In summer of 2022, we became interested in more broadly characterizing enterovirus circulation in Arizona, prompting the use of a pan-enterovirus amplicon assay followed by sequencing, reference database development, and the novel application of QIIME2 [19] for enterovirus characterization. The assay was applied to human nasal and wastewater samples collected from April through October of 2022, and given the increased mixing of individuals at this time in the COVID-19 pandemic, we expected seasonal circulation of several clinically relevant enteroviruses. Of the QIIME2 classified sequencing reads, CV-A6 was most abundant across location types, and was found in samples collected during April–October of 2022. CV-A6 is a more recently identified cause of hand, foot, and mouth disease, and is generally thought to emerge in the late summer to early fall [9]. Similarly, CV-A19, most commonly associated with gastroenteritis and herpangina (i.e. mouth blisters), was detected during April–October of 2022. CV-A4, another enterovirus frequently associated with hand, foot, and mouth disease, was found sporadically in the larger community samples collected from the cities of Flagstaff and Tempe from June through September. The lack of CV-A4 in congregate living sites of Flagstaff may be associated with the younger age range of the population, generally spanning from 18 to 25 years. Surprisingly, we also detected EV-D68 with the pan-enterovirus assay, first just as a one-time low-level hit in June of 2022, but frequently from August to mid-October in both the cities of Flagstaff and Tempe. We determined that detections using the pan-enterovirus versus previous RT-qPCR assays can be attributed to mutations accumulated in primer binding sites during 2018–2022.

Given the discovery of cryptic EV-D68 circulation using the pan-enterovirus assay, we retested all human and wastewater samples from June–October of 2022 using both the CDC2022 and newly developed D68-Detect assays. While we did not detect EV-D68 in human nasal samples, both assays detected the virus in the wastewater from Tempe and Flagstaff, Arizona in early August 2022. Viral load peaked the last week of August and subsided in mid to late October. Trends found in this study are in alignment with expectations of EV-D68 seasonality observed in even years within the United States (Fig. 1), and align with human clinical case trends reported for Phoenix [52,53], Kansas City, MO [54], and the state of Maryland [48]. An additional effort surveyed for EV-D68 in San Jose and Oceanside, CA wastewater, and similarly found evidence of circulation from July 2022 into December 2022, with wastewater viral load trends correlating with counts of human EV-D68 cases in nearby counties [55]. The detection of EV-D68 in both California and Arizona wastewater sewersheds in the latter half of 2022 indicates that even in years when AFM caseloads are low, wastewater-based surveillance can provide an estimate of risk.

The Arizona Department of Health Services reported one case of AFM within the state, occurring in August of 2022 (personal communication and [15]). However, EV-D68 was not detected in cerebrospinal fluid, stool, serum, or nasopharyngeal samples (NP) collected from the patient and tested with RT-qPCR (all samples) or culture (stool). This is not particularly surprising given that in 2018, 2019, and 2020, entero- or rhinoviral RNA was only detected in 50%, 37%, and 26% of AFM cases, respectively, across the United States [56]. Additionally, members of our study team previously found EV-D68 RNA in four of six NP swabs and none of the eleven CSF samples collected from suspect AFM cases in a 2016 Phoenix, AZ pediatric cluster[53]. EV-D68 is not a notifiable disease unless AFM is reported and associated samples are also EV-D68 positive. Otherwise, most clinical systems test for the presence of all enteroviruses and rhinoviruses, and we’ve shown here that a diverse distribution exists at any given time (Fig. 2). This effort demonstrates the utility of wastewater surveillance for the identification of EV-D68 in a population, even in the absence of AFM cases, and explains the increase of enteroviruses and rhinoviruses in the clinical setting (Fig. 4). While the timelines between the EV-D68 wastewater signal and AFM case do coincide, the single patient with AFM in Arizona may have been infected with a different virus that can cause AFM (e.g. flaviviruses, adenoviruses, herpesviruses, and other enteroviruses) [16]. Another possibility is that clearance of EV-D68 viral particles by neutralizing antibodies may have occurred prior to tests being conducted [57]. Our analyses in this study indicate that wastewater-based epidemiology can be a complementary tool to further inform population-level situational awareness to public health agencies and clinicians regarding the presence of EV-D68.

To better understand how EV-D68 genomes from Arizona are related to globally sampled counterparts, we designed a new EV-D68 tiled amplicon sequencing panel using a B3 subclade reference genome. As expected, breadth of coverage was highest for the B3 subclade positive control (∼98%) but was still approximately 86% for the subclade D positive control (Table 1), indicating that this primer scheme should be effective for amplifying across the distribution of EV-D68 when given enough space on a sequencing run. We built a maximum likelihood phylogeny using 31 Arizona genomes sequenced as part of this effort and 934 publicly available genomes. Twenty-nine of the Arizona genomes clustered monophyletically within genomes from Maryland, USA, which were the only other United States-based genomes sequenced in summer and fall of 2022 at the time of submission. The monophyletic cluster indicates a single primary entrance of EV-D68 into Arizona which then circulated throughout the northern and central parts of the state. Given the limited representation of EV-D68 genomes from the United States, it is highly likely that there were many intermediary locations between Maryland and Arizona, rather than a direct link between the two states. Additional sequencing of clinical and wastewater samples from other locations may reveal common trends of EV-D68 dispersal each year, for example, from the eastern to western coasts of the United States. Our hope is that the new EV-D68 tiled amplicon sequencing panel can facilitate additional sequencing efforts.

Our understanding of communicable disease circulation is based primarily on those cases that are reported to public health agencies, information gleaned from syndromic surveillance, and administrative hospital discharge databases. Yet in just the past few years, there have been nationwide and global instances of morbidity and mortality due to non-reportable pathogens known to seasonally circulate (e.g. multiple adenoviruses and adeno-associated virus [58], EV-D68 [11,59,60]). Wastewater testing as an early warning system for viral outbreaks has been suggested by several teams, starting in the early 1940’s [61], but very purposefully implemented during the ongoing COVID-19 pandemic. Our efforts here have demonstrated another use case for wastewater-based epidemiology, as EV-D68 can be routinely screened for and sequenced from wastewater samples. Furthermore, although far fewer AFM cases presented in 2022 than in comparably higher years, RT-qPCR assays deployed in this study and others [44,48,49,50,51] still detected shedding in wastewater. We expect that during larger outbreak years (e.g. 2014, 2016, and 2018), there will be substantially more shedding of EV-D68 into wastewater, and that these approaches will be useful for outbreak monitoring in the absence of EV-D68-specific clinical testing. This research supports the importance of continued wastewater-based surveillance in some key locations over the long-term, to learn more about the magnitude of EV-D68 viral loads and the association with AFM cases in peak years.

## DATA AVAILABILITY STATEMENT

All data produced in the present study are available upon reasonable request to the authors. Sequences from this study are being deposited into NCBI bioproject PRJNA1035117.

## ACKNOWLEDGEMENTS

We thank the human participants that provided samples as part of the IRB approved process.

## AUTHOR CONTRIBUTIONS

## ETHICAL APPROVAL

Institutional Review Board of Northern Arizona University gave ethical approval for this work (Project 1766728).

Institutional Review Board of Northern Arizona Healthcare gave ethical approval for this work (2023-014-FMC).

