## Supplementary figures and images for "Pan-Enterovirus Characterization Reveals Cryptic Circulation of Clinically Relevant Subtypes in Arizona Wastewater"

### Supplemental Figure 1

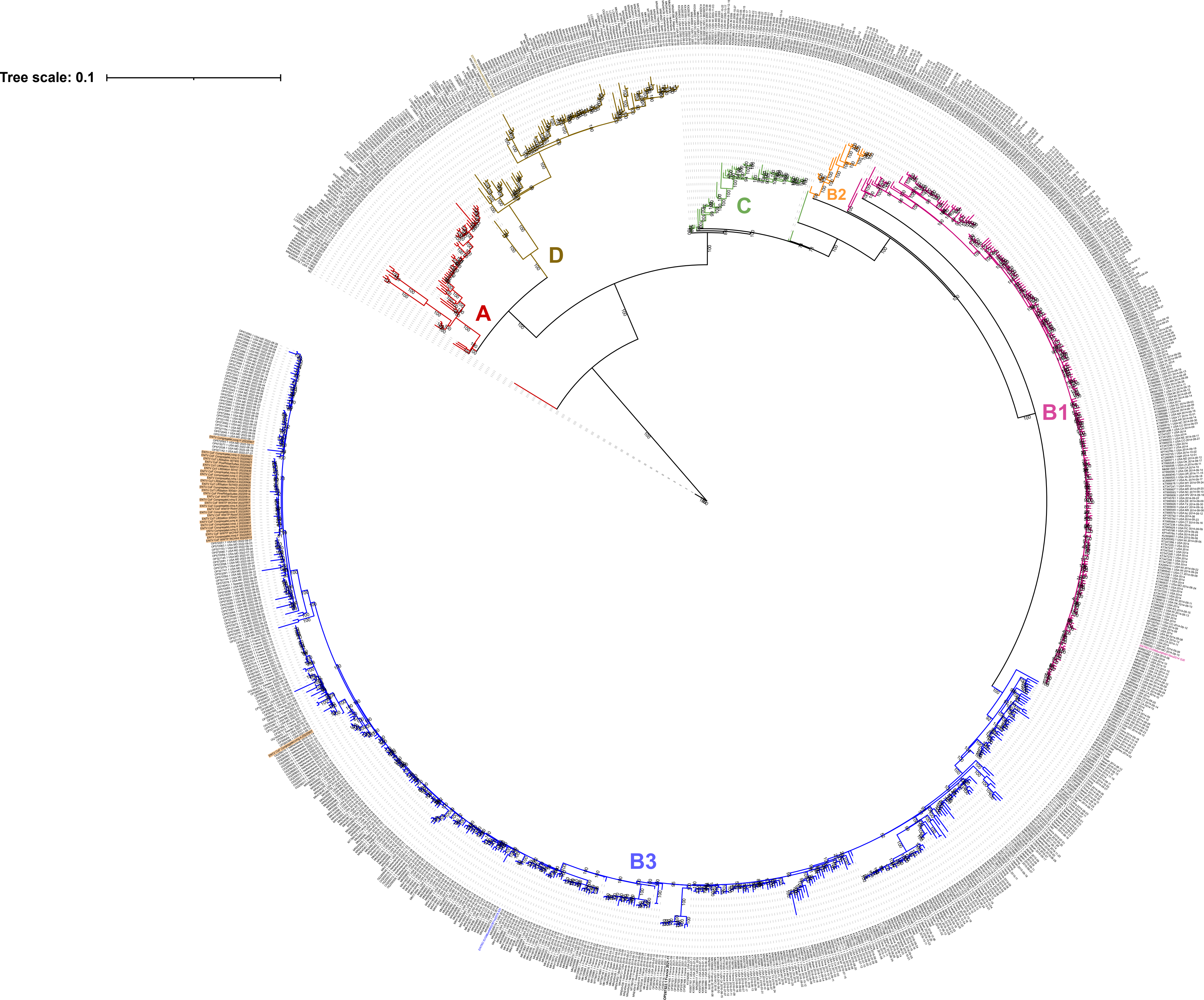
