## Supplemental Table 1 for "Pan-Enterovirus Characterization Reveals Cryptic Circulation of Clinically Relevant Subtypes in Arizona Wastewater"

**Supplemental Table 1:** Droplet Digital PCR (ddPCR) summary results from the 2C Region assay for each BEI Resources (NR-52357) EV-D68 Isolate cDNA dilution, including Reverse Transcription (RT) and ddPCR No Template Controls (NTC) from Direct Quantification Experiment on a BioRad QX200 AutoDG Droplet Digital PCR System. Each sample had greater than 10,000 droplets generated and both NTCs contained zero positive droplets. Manual thresholding was performed on all dilutions with the use of the NTC amplitudes of both NTCs present on the run, the final threshold was 617.56. All dilutions of the EV-D68 Isolate cDNA dilution had positive droplets in the reactions, however, the 1:10,000,000 dilution had zero positive droplets. The 1:10,000,000 dilution was too dilute for ddPCR quantification and detection.

| Sample | Droplet Count | Positive Droplet Count | Negative Droplet Count | Dilution Factor | Threshold | Conc. (copies/µL) | Conc. (copies/Reaction) | Stock Dilution Conc. (copies/µL) | Original Stock Conc. (copies/µL) |
| --- | --- | --- | --- | --- | --- | --- | --- | --- | --- |
| RT_NTC | 17125 | 0 | 17125 | 0 | 617.56 | 0.00 | 0.00 | 0.00 | 0.00 |
| NR-52357  1:100 | 15869 | 15863 | 6 | 0.01 | 617.56 | 9271.02 | 203962.34 | 101981.17 | 10198117.19 |
| NR-52357  1:1,000 | 16701 | 9008 | 7693 | 0.001 | 617.56 | 911.95 | 20062.91 | 10031.45 | 10031453.49 |
| NR-52357  1:10,000 | 16860 | 1334 | 15526 | 0.0001 | 617.56 | 96.97 | 2133.43 | 1066.71 | 10667141.57 |
| NR-52357  1:100,000 | 18086 | 153 | 17933 | 0.00001 | 617.56 | 9.99 | 219.89 | 109.94 | 10994263.84 |
| NR-52357  1:1,000,000 | 17344 | 17 | 17327 | 0.000001 | 617.56 | 1.15 | 25.38 | 12.69 | 12690722.22 |
| NR-52357  1:10,000,000 | 17885 | 0 | 17885 | 0.0000001 | 617.56 | 0.00 | 0.00 | 0.00 | 0.00 |
| ddPCR_NTC | 17281 | 0 | 17281 | 0 | 617.56 | 0.00 | 0.00 | 0.00 | 0.00 |

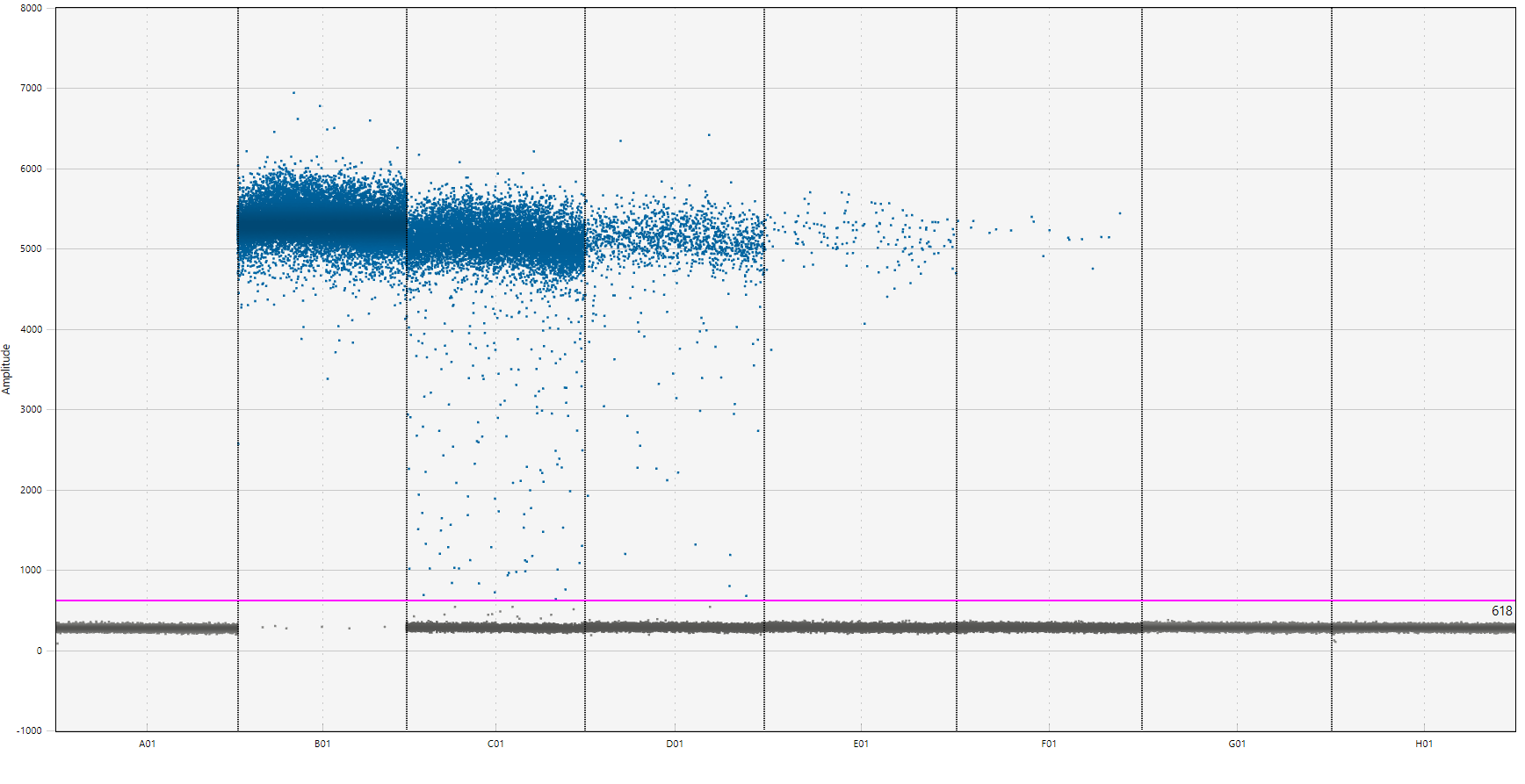

**Supplemental Figure 2a:** Droplet Digital PCR Amplitude plot (y-axis) of 2C Region assay on BEI Resources (NR-52357) EV-D68 isolate cDNA in 10-fold serial dilution (x-axis) from Direct Quantification Experiment on a BioRad QX200 AutoDG Droplet Digital PCR System. The threshold (pink) separates positive droplets (blue) from negative droplets (gray). The Reverse-Transcription No Template Control (A01) and the ddPCR No Template Control (H01) have 100% negative droplets. The No Template Control wells were used to threshold all wells to normalize positive droplets. The dilutions 1:100 (B01), 1:1,000 (C01), 1:10,000 (D01), 1:100,000 (E01), and 1:1,000,000 (F01) all contained positive droplets. The 1:10,000,000 dilution in the series did not contain positive droplets, therefore, it was too dilute for the assay to detect the EV-D68 control.

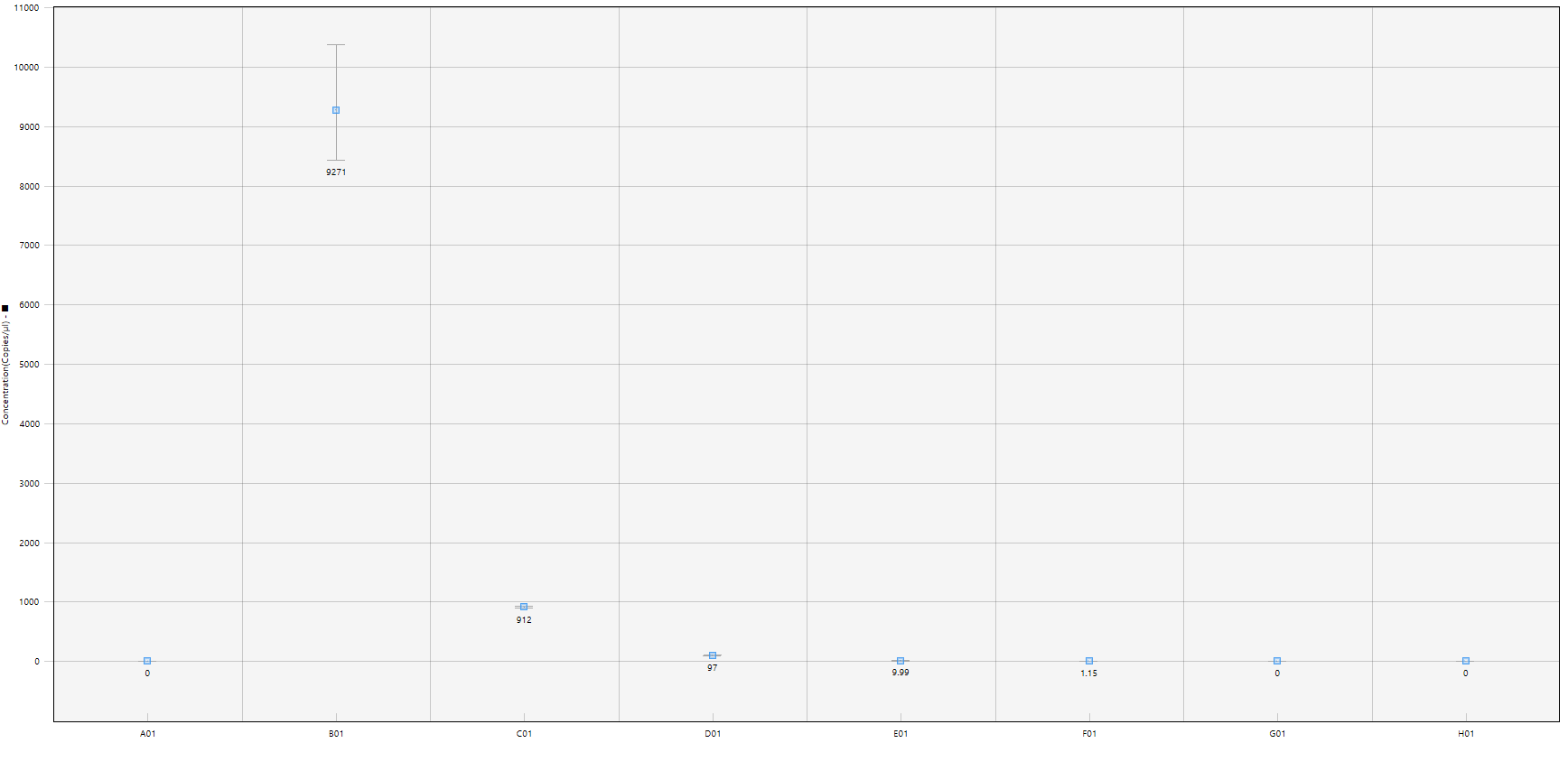

**Supplemental Figure 2b**: Droplet Digital PCR concentrations in copies/µL (y-axis) of 2C Region assay on BEI Resources (NR-52357) EV-D68 isolate cDNA in 10-fold serial dilution (x-axis) from Direct Quantification Experiment on a BioRad QX200 AutoDG Droplet Digital PCR System. Concentrations for each sample are not back-calculated to the original stock concentration. The Reverse-Transcription No Template Control (A01) and the ddPCR No Template Control (H01) a concentration of 0 copies/µL, which reflects the 0 positive droplets observed in Supplemental Table 1. The 1:100 (B01), 1:1,000 (C01), 1:10,000 (D01), 1:100,000 (E01), and 1:1,000,000 (F01) all have a concentration in 10-fold differences from one another. Since the 1:10,000,000 dilution did not contain positive droplets (Supplemental Figure 1, Supplemental Table 1), therefore its concentration was determined to be 0 copies/µL. The 1:100 dilution has very large error bars around its calculated concentration, this is due to the low number of negative droplets, resulting in lower confidence in the poisson algorithm’s concentration estimation of EV-D68 at this dilution.
